# Divergent causal signatures of the adiposity quadrants on coronary artery disease

**DOI:** 10.64898/2026.09.02.26362109

**Authors:** Jesús Abraham Simón-Campos, Hugo Laviada-Molina, Pedro Campos-Corzo, Enrique Castaño-De la Serna

## Abstract

**Background:** Obesity and leanness lie at opposite ends of the weight spectrum, yet both carry coronary risk when metabolically dysfunctional. A 2025 Commission redefined obesity around organ function rather than body mass, but whether that distinction reflects a difference in causal architecture, and whether the two high-risk phenotypes reach disease by distinct routes, is untested.

**Methods:** We conducted two-sample Mendelian randomization of the four combinations of adiposity and metabolic health against coronary artery disease (CARDIoGRAMplusC4D; 60,801 cases), using published adiposity and insulin-resistance instruments. Thirty mediators were interrogated identically in every quadrant by two-step Mendelian randomization with multivariable adjustment, and the pipeline replicated with an independent instrument sharing five of fifty-three variants.

**Results:** Adiposity with preserved metabolic function was protective (odds ratio 0.28, 95% CI 0.19–0.42), whereas adiposity with metabolic dysfunction (1.90, 1.42–2.55) and leanness with metabolic dysfunction (1.86, 1.52–2.27) conferred indistinguishable risk (ratio of odds ratios 1.02; P = 0.91). The unhealthy obese phenotype acted through triglycerides and diabetes; the unhealthy lean through apolipoprotein B, blood pressure and diabetes, sharing no other conduit. High-density lipoprotein cholesterol and triglycerides collapsed despite being the most strongly instrumented exposures, indicating causal redundancy rather than weak-instrument bias.

**Conclusions:** Coronary risk follows the failure of adipose storage capacity, not its magnitude. The two high-risk phenotypes reach the same endpoint by divergent causal architectures that reproduce in full with an independent instrument, and the metabolically unhealthy lean phenotype carries risk equal to clinical obesity while falling outside a framework that requires excess adiposity to be confirmed first.

**What Is New?:**

- Adiposity with preserved metabolic function lowers coronary risk (odds ratio 0.28), whereas adiposity with metabolic dysfunction raises it (1.90); the distinction between preclinical and clinical obesity therefore reflects a difference in causal direction, not a threshold along fat mass.
- Leanness with metabolic dysfunction carries risk indistinguishable from that of unhealthy obesity (1.86; P = 0.91 for the difference), yet falls outside a diagnostic framework that requires excess adiposity to be confirmed before organ function is assessed.
- The two high-risk phenotypes reach coronary disease by divergent routes — triglycerides and diabetes in the obese, apolipoprotein B, blood pressure and diabetes in the lean — converging only on diabetes.

**What Are the Clinical Implications?:**

- The lean patient at coronary risk is not identified by body mass index or by the standard lipid panel; apolipoprotein B and blood pressure are the measures that detect this phenotype.
- Because the causal levers differ between phenotypes, a single risk-mitigation strategy cannot serve both: triglyceride and glycaemic control in the obese phenotype, apolipoprotein B lowering and blood pressure management in the lean.
- Functional assessment of cardiometabolic risk should not be gated behind an anthropometric threshold, since storage failure can occur at any body size.

## Introduction

Obesity is among the most firmly established risk factors for coronary artery disease (CAD), yet the relationship between body weight and cardiovascular risk is neither uniform nor simple. Some individuals carry substantial fat mass with a preserved metabolic profile, while others develop insulin resistance, dyslipidaemia and hypertension at a normal body mass index (BMI). Clinical practice has begun to follow. A 2025 Commission endorsed by more than 75 international medical organisations redefined obesity around the function of tissues and organs rather than around body mass. It distinguishes clinical obesity, in which excess adiposity impairs organ function, from preclinical obesity, in which function is preserved, and demotes BMI to a screening instrument unsuitable as an individual measure of health.^1^ That framework rests on clinical and consensus grounds; whether the distinction it draws corresponds to a difference in causal architecture, rather than to a gradient of risk along fat mass, has not been established.^2,3^ A substantial fraction of individuals living with obesity remain free of metabolic abnormalities, while a comparable fraction of lean individuals harbour the dyslipidaemia, insulin resistance, and hypertension that ordinarily accompany excess weight.^4,5^ This dissociation—captured clinically in the phenotypes of the “metabolically healthy obese” and the “metabolically obese normal-weight” individual—implies that it is the metabolic consequences of adiposity, rather than adiposity itself, that drive cardiovascular risk.^6,7^ The predominant mechanistic explanation is the adipose expandability hypothesis: when subcutaneous adipose tissue reaches the limit of its capacity to store surplus energy safely, lipid is deposited ectopically in liver, muscle, and viscera, precipitating insulin resistance and its cardiometabolic sequelae irrespective of total fat mass.^8–10^

Human genetics has made this framework tractable to causal analysis through Mendelian randomization (MR), which uses genetic variants as instruments to estimate causal effects free of the confounding and reverse causation that limit observational studies. Human genetics offers a way to test this directly. Variants that raise adiposity while conferring a favourable metabolic profile (favourable adiposity) and those that raise it with an adverse profile (unfavourable adiposity) exert opposing causal effects on CAD, disentangling the metabolic from the mechanical consequences of excess weight.^2,4,11^ In parallel, variants marking impaired peripheral fat storage define a lean, insulin-resistant phenotype that carries increased risk of diabetes, hypertension, and coronary disease despite lower body mass index (BMI).^3,5^ Together these instruments render the four quadrants of the adiposity–metabolic health plane—healthy and unhealthy, obese and lean—accessible to causal analysis.^12,13^

To date, however, these quadrants have been studied in isolation. No study has placed all four within a single causal framework. The obese poles have been characterized with favourable and unfavourable adiposity instruments;^2,4,11^ the metabolically unhealthy lean pole with fasting-insulin instruments;^5^ and the mediators of the body-mass-index–to–coronary-disease relationship—chiefly triglycerides and glycaemic traits—have been quantified for the obese phenotype alone.^14^ What has not been established is whether the phenotypes that converge on coronary disease do so through a shared causal pathway or through distinct ones. If the lean and obese phenotypes reach the same endpoint by different biological routes, then the risk of one cannot be captured by the mediators of the other, and a single, weight-centred model of cardiovascular risk assessment would systematically misclassify a substantial population. A second gap follows directly from the new framework. Its diagnostic algorithm requires confirmation of excess adiposity — by BMI, waist circumference or waist-to-height ratio — before organ function is assessed; adiposity remains the gateway to diagnosis.^1^ A phenotype that is metabolically unhealthy without excess adiposity therefore does not enter the framework at all, and is classified as neither clinical nor preclinical obesity. Whether such a phenotype carries risk comparable to clinical obesity is an empirical question with immediate implications for who is screened.

Here we place all four quadrants of the adiposity–metabolic health plane within a single causal framework, instrumented against a common coronary outcome and interrogated with an identical panel of thirty candidate mediators. We estimate the total causal effect of each quadrant, dissect the mediating conduits through which each acts, and test whether the risk architectures of the phenotypes at comparable risk are shared or divergent. We further replicate the metabolically unhealthy lean axis—both its total effect and its mediating signature—using an independent instrument constructed nearly a decade later by different methods,^7^ to establish whether any observed signature is a robust feature of the biology or an artefact of instrument choice.

## Methods

### Study design

We conducted a two-sample MR study to test the causal roles of the four quadrants of the adiposity–metabolic health plane (metabolically healthy or unhealthy × obese or lean) on CAD, and to dissect the causal conduits through which each quadrant acts. We report the study in accordance with the STROBE-MR guidelines.^40^ MR rests on three assumptions: that the genetic instruments are robustly associated with the exposure (relevance), share no common cause with the outcome (independence), and affect the outcome only through the exposure (exclusion restriction).^41^ We evaluated these assumptions through the comprehensive sensitivity framework described below. All analyses used publicly available genome-wide association study summary statistics from studies that obtained appropriate ethical approval and participant consent; no new ethical approval was required. We restricted analyses to participants of European ancestry to minimize population stratification.

### Genetic instruments

#### Favourable and unfavourable adiposity (obese quadrants)

All exposure instruments and outcome datasets, with their sources, accessions and sample sizes, are listed in Table 1. To instrument the two obese quadrants, we used the metabolically favourable and unfavourable adiposity variants reported by Martin et al.,^2^ derived from a multivariate genome-wide association study (GWAS) of body fat percentage together with a composite metabolic phenotype in UK Biobank, in which the two clusters were separated by k-means clustering on high-density lipoprotein cholesterol, sex hormone-binding globulin, triglycerides and liver enzymes. Blood pressure did not enter the derivation of either cluster. ^2021;11:^ Of the 36 favourable and 38 unfavourable adiposity variants originally reported, 34 and 27 respectively were available after harmonization against the outcome; the remainder were lost through absence from the outcome dataset, ambiguous palindromic alleles, or lack of a suitable proxy.

**Table 1.** Genetic instruments and outcome datasets. Source, number of variants, sample size, and accession identifier for each exposure instrument and outcome dataset. Case and control counts are given for binary outcomes; n/a indicates that the field does not apply (outcome datasets are not instrumented). The insulin resistance variant set was taken from Supplementary Table 3 of Lotta et al., with effect sizes from the fasting insulin adjusted for body mass index genome-wide association study (ebi-a-GCST007857); the multivariate insulin resistance variant set was taken from the supplementary material of Ye et al. The two lean rows use the same 53-variant instrument, with the effect allele inverted for the healthy pole. Accession ebi-a-GCST011364 corresponds to the UK Biobank component of the Hartiala 2021 myocardial infarction study (17,505 cases, 454,212 controls), not to the full trans-ancestry meta-analysis; it is therefore independent of CARDIoGRAMplusC4D. The FinnGen phenotype is registered as “major coronary heart disease event” (finn-b-I9_CHD, release R5). ᵃ The favourable and unfavourable adiposity variants were identified by Martin et al. (Diabetes 70, 1843–1856, 2021); the effect size estimates used here are those of the body fat percentage genome-wide association study in 442,278 European-ancestry UK Biobank participants reported by Ji et al., from which that sample size derives. *Abbreviations used in this section: ApoB, apolipoprotein B; BMI, body mass index; CAD, coronary artery disease; CI, confidence interval; FA, favourable adiposity; GWAS, genome-wide association study; HDL, high-density lipoprotein; IVW, inverse-variance weighted; MR, Mendelian randomization; MVMR, multivariable Mendelian randomization; mvIR, multivariate insulin resistance; OR, odds ratio; SBP, systolic blood pressure; T2D, type 2 diabetes; UFA, unfavourable adiposity; VLDL, very-low-density lipoprotein.*

| Role | Trait | Source | n var. | Sample (cases/controls) | Accession |
| --- | --- | --- | --- | --- | --- |
| Exposure | Favourable adiposity | Martin 2021 <sup>a</sup> | 34 | 442,278 | ukb-b-8909 |
| Exposure | Unfavourable adiposity | Martin 2021 <sup>a</sup> | 27 | 442,278 | ukb-b-8909 |
| Exposure | Insulin resistance | Lotta 2017 | 53 | 108,557 | ebi-a-GCST007857 |
| Exposure | Insulin resistance (inverted) | Lotta 2017 | 53 | 108,557 | ebi-a-GCST007857 |
| Exposure | Multivariate insulin resistance | Ye 2025 | 351 | ≈400,000 | Ye 2025, Suppl. |
| Outcome | CAD (primary) | Nikpay 2015 | n/a | 60,801 / 123,504 | ieu-a-7 |
| Outcome | Myocardial infarction | Hartiala 2021 | n/a | 17,505 / 454,212 | ebi-a-GCST011364 |
| Outcome | Coronary heart disease | FinnGen R5 | n/a | 21,012 / 197,780 | finn-b-I9_CHD |

#### Insulin resistance (unhealthy lean quadrant)

We instrumented the metabolically unhealthy lean quadrant using the 53 insulin resistance variants reported by Lotta et al.,^5^ defined by their association with fasting insulin adjusted for BMI and a concordant pattern of lower high-density lipoprotein cholesterol and higher triglycerides. Where standard errors were not directly tabulated, we reconstructed them from the reported effect estimates and exact P values using the standard normal quantile relationship.

#### Healthy lean quadrant

The metabolically healthy lean quadrant represents the protective pole of the same insulin-sensitivity axis. We examined it as a directional consistency analysis, applying the Lotta instrument with the sign of the effect allele inverted; the resulting estimates are the exact mirror of the unhealthy lean quadrant.

#### Replication instrument

To test whether findings depended on the choice of instrument, we independently replicated the lean axis using the multivariate insulin resistance instrument reported by Ye et al.,^7^ comprising 351 variants derived from a multivariate genome-wide analysis of insulin resistance phenotypes. Beyond reproducing the total effect and the individual step-a associations, we repeated the entire analytical pipeline with this instrument alone: the same 30-mediator panel was screened, mediators were selected by Benjamini-Hochberg false discovery rate rather than by a nominal threshold, and the selected set was carried into multivariable Mendelian randomization. This provides a replication of the analytical procedure and not only of its output. Conditional F-statistics and Cochran’s Q for the multivariable model are reported in Tables S1 and S2; conditional F-statistics assume zero genetic covariance between exposures, which is conservative with respect to instrument strength.

### Outcome data

Our primary outcome was CAD from the CARDIoGRAMplusC4D consortium reported by Nikpay et al., ^2021;42:^ comprising 60,801 cases and 123,504 controls, accessed through the IEU OpenGWAS database.^43^ We replicated the metabolically unhealthy lean axis against myocardial infarction from the UK Biobank component of the study of Hartiala et al. (17,505 cases and 454,212 controls, accession ebi-a-GCST011364), which does not overlap CARDIoGRAMplusC4D^44^ and coronary heart disease from the FinnGen study.^45^

### Mediators and mediation analysis

To dissect the causal conduits linking each quadrant to CAD, we assembled a harmonized panel of 30 candidate mediators spanning lipids and lipoproteins, blood pressure, glycaemic traits, inflammatory and adipokine markers, ectopic fat, branched-chain amino acids, renal function, serum urate, and behavioural exposures (Table S3).

Blood pressure was taken from GWAS not adjusted for BMI, because summary statistics conditioned on BMI introduce collider bias when the exposure is itself a measure of adiposity. We assessed mediation using a two-step MR framework.^39,46^ We considered a mediator an active conduit only when both steps were directionally consistent and statistically supported. Because individual mediation proportions estimated by the product-of-coefficients method are inflated and non-additive in the presence of correlation among conduits, we did not report point estimates of the total mediated proportion. Instead, we established the independent contribution of each conduit through multivariable MR (MVMR), modelling candidate conduits jointly and estimating their conditional direct effects, with instrument strength quantified by the conditional F-statistic.

### Statistical analysis

We obtained the primary causal estimates by the multiplicative random-effects IVW method,^47^ and evaluated horizontal pleiotropy with the MR-Egger intercept test,^48^ using the MR-Egger slope and the weighted median estimator as complementary estimates.^48,49^ We applied MR-PRESSO to detect and correct for outlying variants,^50^ quantified between-variant heterogeneity with Cochran’s Q, and confirmed the direction of each causal relationship with the Steiger test. We performed all analyses in R using the TwoSampleMR, MVMR, and MR-PRESSO packages, querying summary statistics through the IEU OpenGWAS database. The full analysis code, including every model reported here, is deposited at Zenodo (DOI: 10.5281/zenodo.21682531).^43^

### Data availability

All derived data, result tables, figures, and analysis code are openly available at Zenodo (DOI: 10.5281/zenodo.21682531). The analyses used publicly available genome-wide association summary statistics; primary sources are cited in the text and detailed in Table S4.

### Code availability

All analysis code is openly available at Zenodo (DOI: 10.5281/zenodo.21682531). Analyses were performed in R using the TwoSampleMR, MVMR and MR-PRESSO packages.

## Results

### Risk follows metabolic dysfunction, not weight

Across the four quadrants, the causal effect on CAD tracked metabolic health rather than body weight (Figure 1, Table 2). The two metabolically unhealthy quadrants raised risk to a comparable degree despite occupying opposite extremes of the weight spectrum: unfavourable adiposity (unhealthy obese) conferred an odds ratio (OR) of 1.90 (95% confidence interval (CI) 1.42–2.55, P = 1.8×10⁻⁵), and genetically instrumented insulin resistance (unhealthy lean) an OR of 1.86 (1.52–2.27, P = 1.6×10⁻⁹). Conversely, the two metabolically healthy quadrants lowered risk: favourable adiposity (healthy obese) yielded an OR of 0.28 (0.19–0.42, P = 1.1×10⁻⁹), and the healthy lean pole, examined as a directional consistency analysis, the mirror-image estimate of 0.54 (0.44–0.66, P = 1.6×10⁻⁹). Inverse-variance weighted (IVW) and weighted median estimates were closely concordant in all four quadrants (Figure 2). MR-Egger was substantially less precise and did not reach significance in any quadrant; in the healthy obese quadrant the MR-Egger estimate was null (OR 0.91, 95% CI 0.24–3.43, P = 0.90) despite a strongly protective primary estimate (Table 2). This pattern indicates that coronary risk follows adipose storage dysfunction rather than adiposity itself.

**Figure 1.**
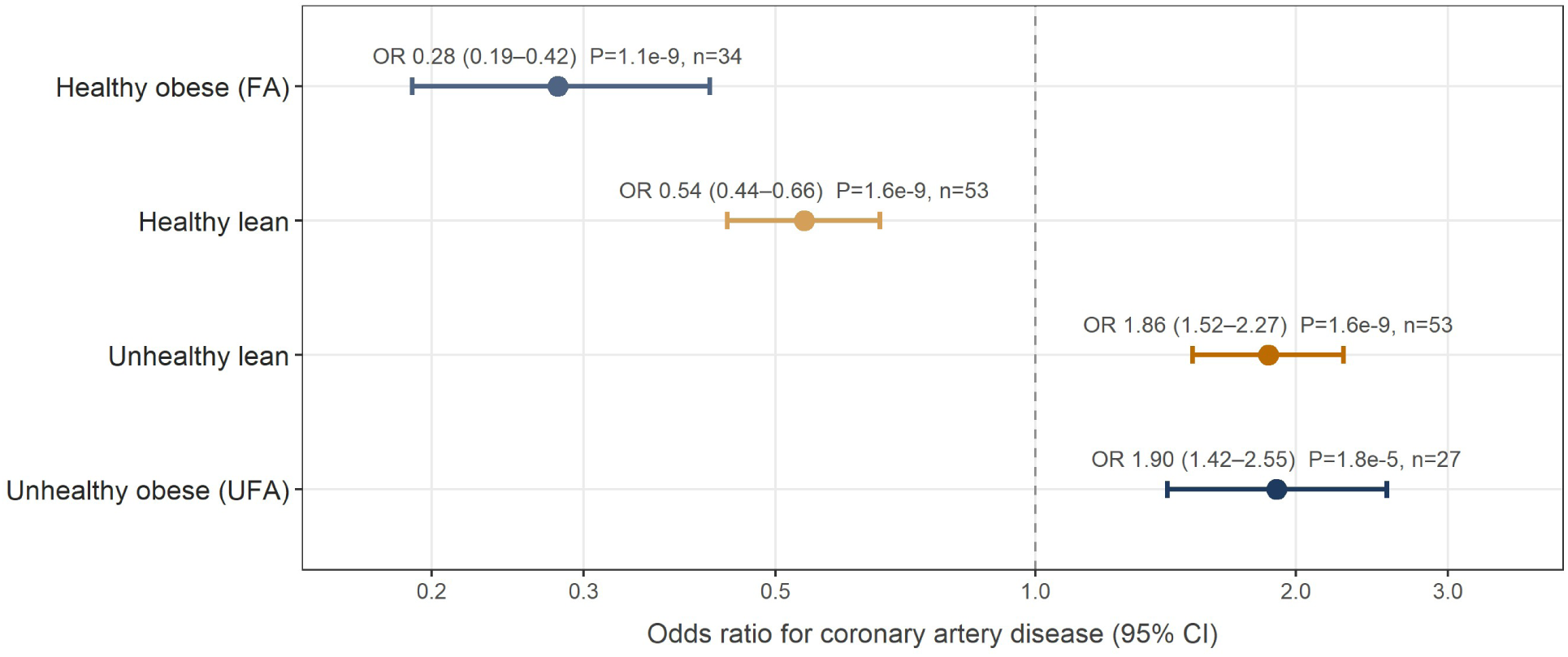
Total causal effects of the four adiposity–metabolic health quadrants on coronary artery disease. Odds ratios with 95% confidence intervals from two-sample inverse-variance weighted Mendelian randomization against CARDIoGRAMplusC4D (60,801 cases, 123,504 controls); n denotes the number of genetic variants retained after harmonization, and P values are two-sided and unadjusted. Colour denotes the quadrant: dark blue, obese poles; ochre, lean poles, with the lighter shade of each pair marking the metabolically healthy quadrant. The two metabolically unhealthy poles confer indistinguishable risk despite opposite body weight (ratio of odds ratios 1.02, 95% CI 0.72– 1.46; P = 0.91 for the difference), whereas among the protective poles favourable adiposity is the stronger (ratio of odds ratios 0.52, 95% CI 0.33–0.81; P = 0.004). The healthy lean quadrant is the mirror of the unhealthy lean quadrant — the same 53-variant instrument with the effect allele inverted — and is shown to complete the 2×2 architecture rather than as independent evidence. Effects are expressed per unit of each instrument; because the adiposity and insulin-resistance instruments are scaled to different exposures, the similarity between the unhealthy poles should be read as concordance of direction and magnitude of risk, not as biologically equivalent increments of exposure. *Abbreviations used in this section: ApoB, apolipoprotein B; BMI, body mass index; CAD, coronary artery disease; CI, confidence interval; FA, favourable adiposity; GWAS, genome-wide association study; HDL, high-density lipoprotein; IVW, inverse-variance weighted; MR, Mendelian randomization; MVMR, multivariable Mendelian randomization; mvIR, multivariate insulin resistance; OR, odds ratio; SBP, systolic blood pressure; T2D, type 2 diabetes; UFA, unfavourable adiposity; VLDL, very-low-density lipoprotein.*

**Figure 2.**
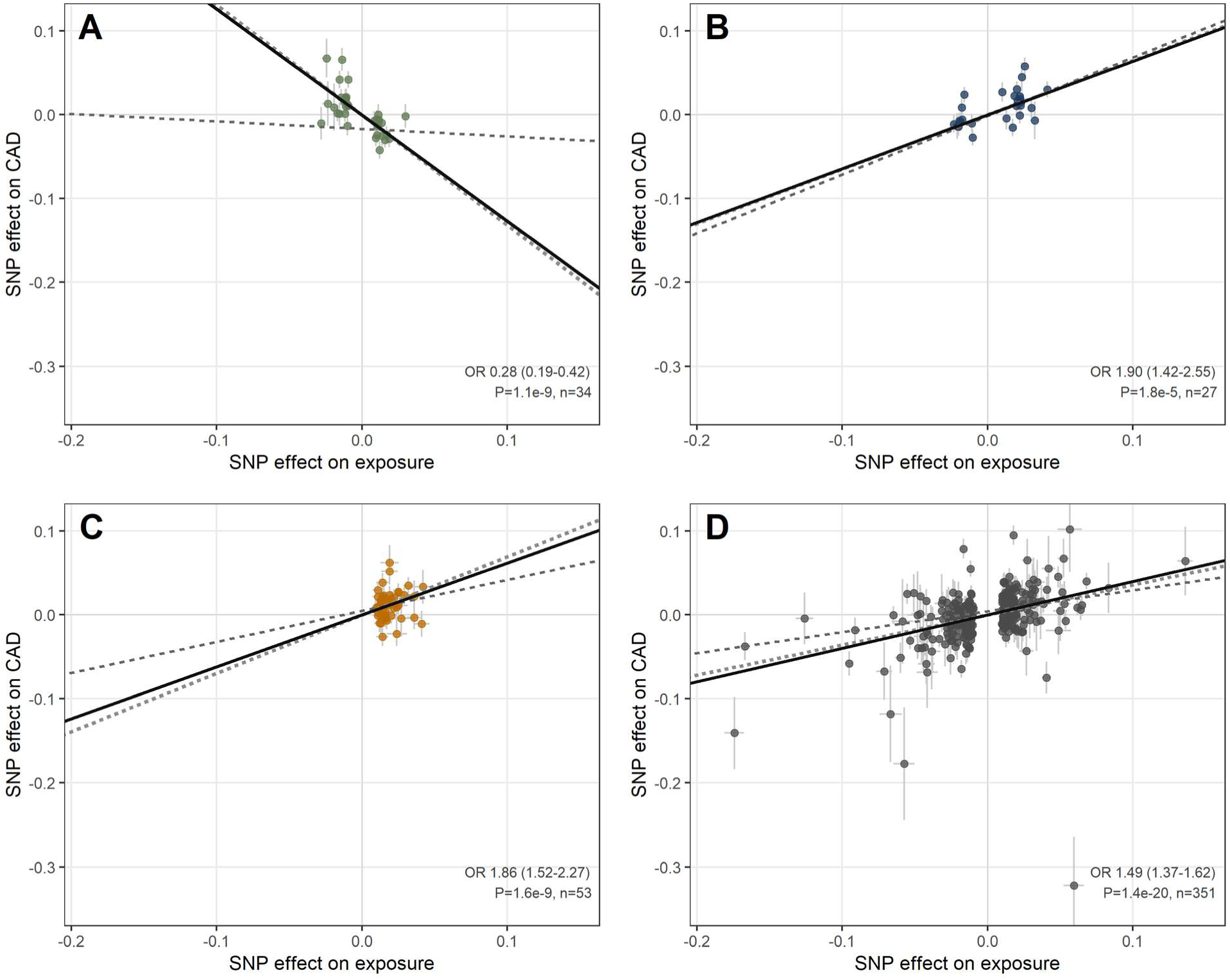
Mendelian randomization scatter plots for the four quadrants against coronary artery disease, drawn on uniform axes to allow direct comparison of slopes. Each point is a genetic variant, plotted by its effect on the exposure (x axis) and on coronary artery disease (y axis), with 95% confidence intervals; lines show the inverse-variance weighted (solid), MR-Egger (dashed) and weighted median (dotted) estimates. Odds ratio, confidence interval, P value and n are shown in each panel. (A) Healthy obese; (B) unhealthy obese; (C) unhealthy lean (Lotta instrument); (D) unhealthy lean (mvIR instrument). The inverse-variance weighted and weighted median estimates are closely concordant in all four panels. MR-Egger is markedly less precise and does not reach significance in any of the three quadrant panels (A–C); in (A) the MR-Egger slope is essentially null (OR 0.91, 95% CI 0.24–3.43, P = 0.90) despite a strongly negative inverse-variance weighted slope, a divergence expected when the instrument spans a limited range of the exposure and the Egger intercept is imprecisely estimated (intercept P = 0.077). For the mvIR instrument in (D), which comprises 351 variants, the MR-Egger estimate is attenuated but remains significant (OR 1.28, 95% CI 1.11–1.48, P = 7.2×10⁻⁴). Egger intercepts for all four quadrants are reported in Table S5. *Abbreviations used in this section: ApoB, apolipoprotein B; BMI, body mass index; CAD, coronary artery disease; CI, confidence interval; FA, favourable adiposity; GWAS, genome-wide association study; HDL, high-density lipoprotein; IVW, inverse-variance weighted; MR, Mendelian randomization; MVMR, multivariable Mendelian randomization; mvIR, multivariate insulin resistance; OR, odds ratio; SBP, systolic blood pressure; T2D, type 2 diabetes; UFA, unfavourable adiposity; VLDL, very-low-density lipoprotein.*

**Table 2.** Total causal effects of the four adiposity–metabolic health quadrants on coronary artery disease. Odds ratios (95% confidence interval) and P values from inverse-variance weighted (IVW), MR-Egger, and weighted median Mendelian randomization against CARDIoGRAMplusC4D. n, number of genetic variants after harmonization. The healthy lean quadrant is the mirror of the unhealthy lean quadrant (identical instrument, inverted effect allele) and is shown for symmetry; it does not constitute independent evidence. Estimates are directionally concordant across the three methods, with the exception of the healthy obese quadrant, in which the MR-Egger estimate is null (OR 0.91, P = 0.90). All three MR-Egger estimates are non-significant with wide confidence intervals, as expected for instruments of this size; MR-Egger is reported for completeness rather than as a confirmatory estimator. *Abbreviations used in this section: ApoB, apolipoprotein B; BMI, body mass index; CAD, coronary artery disease; CI, confidence interval; FA, favourable adiposity; GWAS, genome-wide association study; HDL, high-density lipoprotein; IVW, inverse-variance weighted; MR, Mendelian randomization; MVMR, multivariable Mendelian randomization; mvIR, multivariate insulin resistance; OR, odds ratio; SBP, systolic blood pressure; T2D, type 2 diabetes; UFA, unfavourable adiposity; VLDL, very-low-density lipoprotein.*

| Quadrant | n | Method | OR (95% CI) | P |
| --- | --- | --- | --- | --- |
| Healthy obese (FA) | 34 | IVW | 0.28 (0.19–0.42) | $1.1 \times 10^{-9}$ |
|  |  | MR-Egger | 0.91 (0.24–3.43) | 0.90 |
| | | Weighted median | 0.27 (0.17–0.43) | $4.4 \times 10^{-8}$ |
| Unhealthy obese (UFA) | 27 | IVW | 1.90 (1.42–2.55) | $1.8 \times 10^{-5}$ |
|  |  | MR-Egger | 2.01 (0.75–5.40) | 0.18 |
| | | Weighted median | 1.91 (1.44–2.55) | $7.8 \times 10^{-6}$ |
| Unhealthy lean (Lotta) | 53 | IVW | 1.86 (1.52–2.27) | $1.6 \times 10^{-9}$ |
|  |  | MR-Egger | 1.45 (0.80–2.61) | 0.22 |
| | | Weighted median | 2.00 (1.56–2.58) | $6.1 \times 10^{-8}$ |
| Healthy lean (Lotta, inverted) | 53 | IVW | 0.54 (0.44–0.66) | $1.6 \times 10^{-9}$ |
|  |  | MR-Egger | 0.69 (0.38–1.25) | 0.22 |
| | | Weighted median | 0.50 (0.38–0.65) | $1.5 \times 10^{-7}$ |

### Divergent, bidirectional causal signatures

Applying an identical panel of 30 mediators to each quadrant, we found that the two unhealthy quadrants, though comparable in the magnitude of their effect, reached CAD through distinct causal conduits (Figure 3, Table 3). In multivariable models restricted to independently identified conduits, the unhealthy obese quadrant acted through a triglyceride–diabetes axis: triglycerides (direct effect β = 0.238, P = 2.0×10⁻⁸) and type 2 diabetes (T2D) (β = 0.138, P = 9.2×10⁻⁸). The unhealthy lean quadrant instead acted through an apolipoprotein B (ApoB)–blood pressure– diabetes axis (Figure 4): ApoB (β = 0.583, P = 3.3×10⁻¹⁶), systolic blood pressure (SBP) (β = 0.037, P = 3.6×10⁻¹⁷), and T2D (β = 0.172, P = 7.0×10⁻⁸).

**Figure 3.**
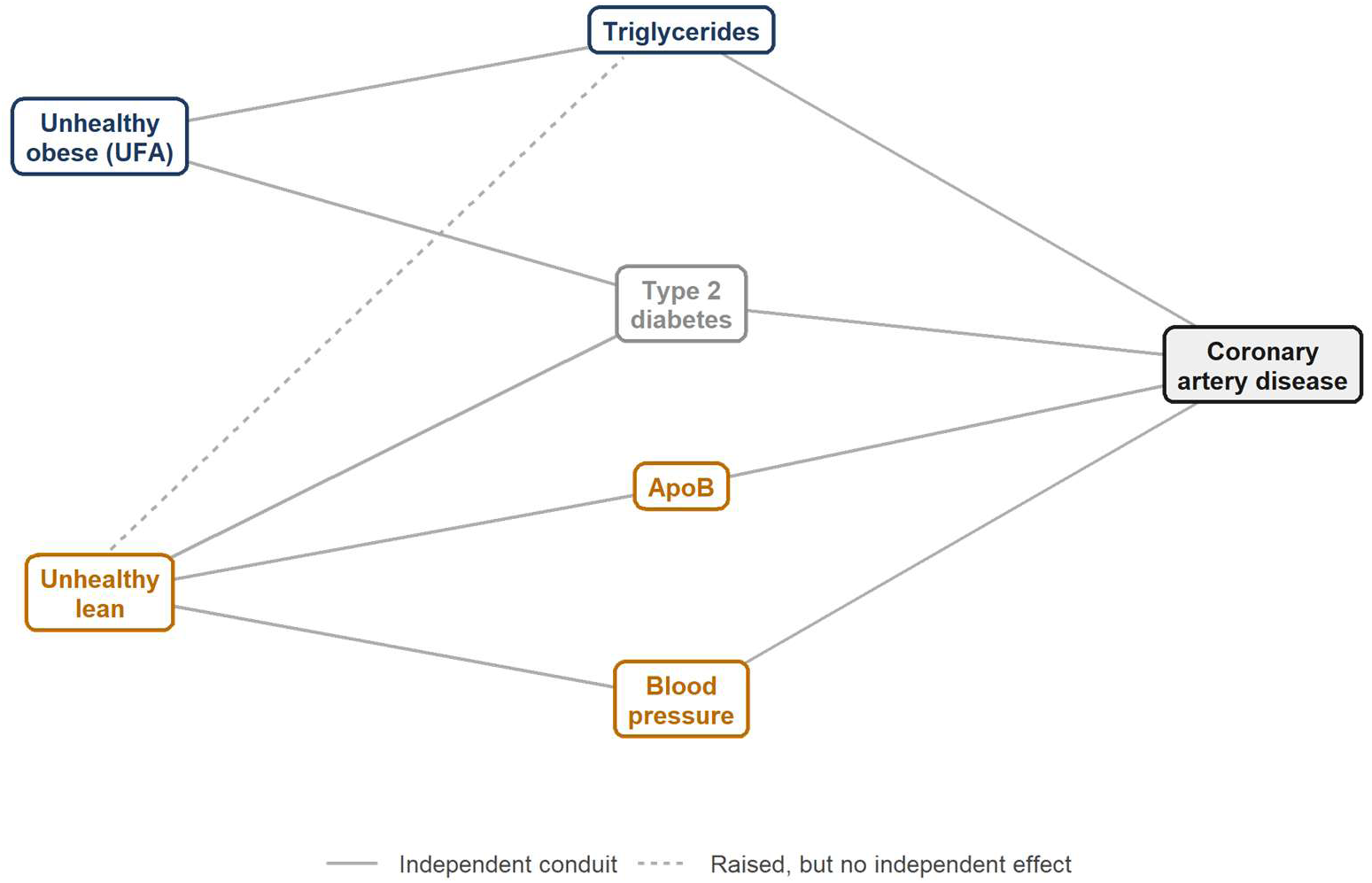
Divergent causal signatures of the two metabolically unhealthy quadrants. Schematic of the conduits linking each unhealthy phenotype to coronary artery disease, derived from two-step Mendelian randomization with multivariable adjustment. The unhealthy obese phenotype (dark blue) acts through triglycerides and type 2 diabetes; the unhealthy lean phenotype (ochre) through apolipoprotein B, blood pressure, and type 2 diabetes. Blood pressure was modelled as systolic blood pressure in the multivariable analysis (Table 3). Type 2 diabetes (grey) is retained in both models. The two exclusions differ in kind: apolipoprotein B is not affected by the unhealthy obese phenotype (β = −0.046, P = 0.66), whereas triglycerides are strongly raised by the unhealthy lean phenotype (dashed connector; β = 1.019, P = 6.3×10⁻¹³) but retain no independent effect on coronary artery disease once apolipoprotein B and blood pressure enter the multivariable model. Solid connectors denote conduits with an independent conditional direct effect; direct effects and conditional F-statistics are given in Table 3. *Abbreviations used in this section: ApoB, apolipoprotein B; BMI, body mass index; CAD, coronary artery disease; CI, confidence interval; FA, favourable adiposity; GWAS, genome-wide association study; HDL, high-density lipoprotein; IVW, inverse-variance weighted; MR, Mendelian randomization; MVMR, multivariable Mendelian randomization; mvIR, multivariate insulin resistance; OR, odds ratio; SBP, systolic blood pressure; T2D, type 2 diabetes; UFA, unfavourable adiposity; VLDL, very-low-density lipoprotein.*

**Figure 4.**
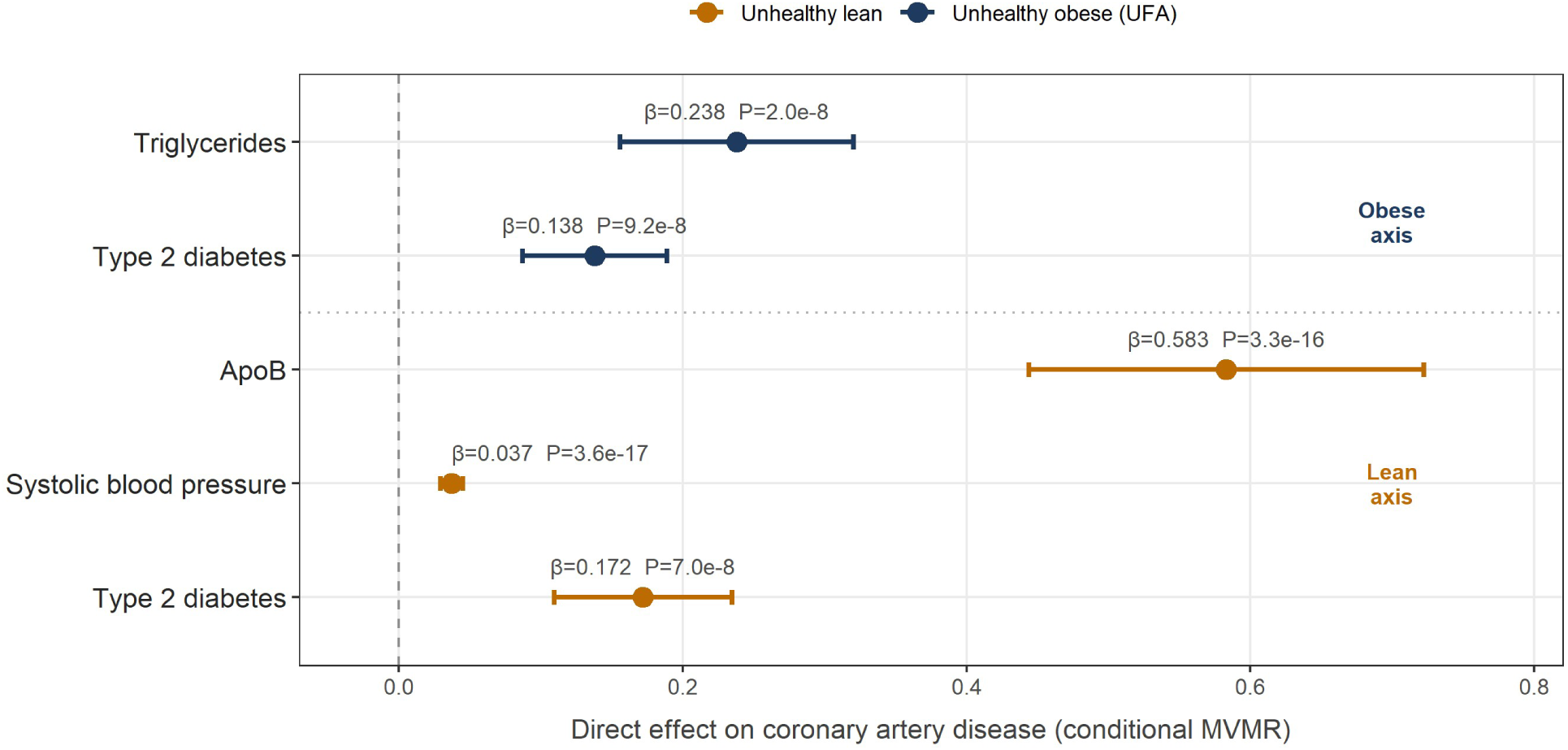
Conditional direct effects of the mediating conduits on coronary artery disease. Direct effects (β) with 95% confidence intervals and P values from multivariable Mendelian randomization; all conditional F-statistics exceed 10 (Table 3). Conduits are grouped by the mediator set nominated by each quadrant at step a: the obese axis (dark blue; triglycerides, type 2 diabetes) above and the lean axis (ochre; apolipoprotein B, systolic blood pressure, type 2 diabetes) below, separated by a dotted line. Estimates are conditional on the mediator set in which each conduit was modelled and are therefore not directly comparable between axes; the axis label denotes the adjustment set, not a stratification of individuals. Type 2 diabetes is retained in both models with closely similar direct effects. *Abbreviations used in this section: ApoB, apolipoprotein B; BMI, body mass index; CAD, coronary artery disease; CI, confidence interval; FA, favourable adiposity; GWAS, genome-wide association study; HDL, high-density lipoprotein; IVW, inverse-variance weighted; MR, Mendelian randomization; MVMR, multivariable Mendelian randomization; mvIR, multivariate insulin resistance; OR, odds ratio; SBP, systolic blood pressure; T2D, type 2 diabetes; UFA, unfavourable adiposity; VLDL, very-low-density lipoprotein.*

**Table 3.** Conditional direct effects of the mediating conduits on coronary artery disease (differential signatures). Direct effect (β), standard error (SE), P value, and conditional F-statistic from multivariable Mendelian randomization for each surviving conduit, by unhealthy quadrant. All conditional F-statistics exceed 10, indicating adequate identification. Direct effects are conditional on the mediator set nominated by each quadrant at step a and are therefore not comparable between quadrants: the “quadrant” label denotes the adjustment set, not a stratification of individuals. Type 2 diabetes is retained in both models. The exclusion of apolipoprotein B from the unhealthy obese axis rests on the absence of a step-a effect (β = −0.046, P = 0.66) and is therefore unconditional; the exclusion of triglycerides from the unhealthy lean axis is conditional, since the lean quadrant does raise triglycerides at step a (β = 1.019, P = 6.3×10⁻¹³) but they retain no independent effect once apolipoprotein B and blood pressure are included. *Abbreviations used in this section: ApoB, apolipoprotein B; BMI, body mass index; CAD, coronary artery disease; CI, confidence interval; FA, favourable adiposity; GWAS, genome-wide association study; HDL, high-density lipoprotein; IVW, inverse-variance weighted; MR, Mendelian randomization; MVMR, multivariable Mendelian randomization; mvIR, multivariate insulin resistance; OR, odds ratio; SBP, systolic blood pressure; T2D, type 2 diabetes; UFA, unfavourable adiposity; VLDL, very-low-density lipoprotein.*

| Quadrant | Conduit | $\beta$ | SE | P | Cond. F |
| --- | --- | --- | --- | --- | --- |
| Unhealthy obese (UFA) | Triglycerides | 0.238 | 0.042 | $2.0 \times 10^{-8}$ | 116.8 |
| Unhealthy obese (UFA) | Type 2 diabetes | 0.138 | 0.026 | $9.2 \times 10^{-8}$ | 31.2 |
| Unhealthy lean (Lotta) | Apolipoprotein B | 0.583 | 0.071 | $3.3 \times 10^{-16}$ | 33.1 |
| Unhealthy lean (Lotta) | Systolic blood pressure | 0.037 | 0.004 | $3.6 \times 10^{-17}$ | 40.9 |
| Unhealthy lean (Lotta) | Type 2 diabetes | 0.172 | 0.032 | $7.0 \times 10^{-8}$ | 14.9 |

*The healthy lean and healthy obese quadrants act through the protective (inverse) direction of the same conduits and are not tabulated separately to avoid redundancy; their mediation mirrors the unhealthy poles.*

The discriminating conduits separated at the exposure-to-mediator step. The lean instrument raised ApoB, SBP and very-low-density lipoprotein cholesterol; the obese instrument moved none of them (β = −0.046, P = 0.66; β = 0.083, P = 0.26; and β = 0.062, P = 0.60 in the obese quadrant), whereas the obese quadrant uniquely elevated leptin, visceral adipose tissue and inflammatory markers. The two axes converged on T2D, which contributed comparably in both (β = 1.599 versus 1.571). Both quadrants raised triglycerides, yet triglycerides retained an independent direct effect on CAD only in the obese multivariable model. This constitutes a divergent, bidirectional signature: two distinct causal routes to the same endpoint, with body weight irrelevant to the distinction.

### Robustness and sensitivity

The total effects were robust across the full sensitivity framework (Table S5). MR-Egger intercepts showed no evidence of directional pleiotropy in any quadrant (P = 0.077, 0.905, and 0.383 for favourable adiposity, unfavourable adiposity, and the lean axis, respectively). The MR-PRESSO global test was significant in all four quadrants. In the two obese quadrants the outlier-corrected estimates were close to the primary estimates (OR 0.30, 95% CI 0.21–0.44; and 1.84, 1.46–2.32). In the lean quadrants the distortion test identified no individual outlying variant after correction for multiple testing, so no outlier-corrected estimate could be computed. The Steiger test confirmed the correct causal direction throughout (P = 2×10⁻⁴⁹, 8×10⁻¹⁵², and 5×10⁻¹³⁶). Although Cochran’s Q indicated between-variant heterogeneity in all quadrants, the concurrently clean MR-Egger intercepts indicate that this heterogeneity was balanced rather than directional—an expected feature of polygenic instruments that does not bias the IVW estimate. Funnel plots were symmetrical about the IVW estimate in every quadrant (Figure S1).

### The healthy obese quadrant reflects transient reserve

The genetic estimate for favourable adiposity must not be mistaken for evidence that obesity is cardioprotective. What the favourable adiposity instrument captures is not protection conferred by fat, but a greater capacity for safe lipid storage in subcutaneous depots. This capacity is finite and temporary: the favourable adiposity instrument was the only one to raise leptin (β = 2.16, P = 6×10⁻¹¹) and the only one that did not reduce visceral adipose tissue (P = 0.85), marking a residual adipose burden invisible to a metabolic instrument. We reasoned that any residual risk detectable within this transient window would have to be carried by a conduit independent of the metabolic profile. Our systematic search identified a single such conduit: lipoprotein(a). Favourable adiposity modestly raised lipoprotein(a) (β = 0.15, P = 0.001), which was a robust causal factor for CAD in the joint model (β = 0.19, P = 2×10⁻¹⁰); no variant of the favourable adiposity instrument lies within 1 Mb of the LPA locus, so this association is not an artefact of instrument composition. Leptin, despite strong elevation, did not causally affect CAD (P = 0.62) and thus functioned as a marker of fat mass rather than a conduit; arterial stiffness, resistin, and N-terminal pro-B-type natriuretic peptide showed no supporting evidence. Beyond lipoprotein(a), no circulating biomarker accounted for residual risk (Table S3).

### Replication with an independent instrument

The metabolically unhealthy lean axis replicated fully with the independent multivariate insulin resistance instrument (351 variants; Figure S2, Tables S6 and S7). This instrument reproduced the total effect on CAD (OR 1.49, 95% CI 1.37–1.62, P = 1.4×10⁻²⁰). It reproduced the mediating signature in its entirety, significantly elevating ApoB (β = 0.315, P = 1×10⁻²³), SBP (β = 0.075, P = 8.2×10⁻⁸), and T2D (β = 0.642, P = 1×10⁻²³)—the identical ApoB–blood pressure–diabetes signature identified with the Lotta instrument. The effect further replicated in two datasets independent of the primary outcome: myocardial infarction in the UK Biobank component of the Hartiala study (OR 2.38, 95% CI 1.86–3.05, P = 5.1×10⁻¹²) and coronary heart disease in FinnGen (OR 2.09, 1.58–2.76, P = 1.9×10⁻⁷). We obtained replication estimates by IVW MR only, and applied the full sensitivity framework to the primary outcome. To test whether the lean signature depended on the analytical pipeline rather than on the biology, we repeated the entire sequence — screening of the 30-mediator panel, selection by false discovery rate, and multivariable modelling — using the mvIR instrument alone. Twenty-four of the thirty mediators passed a 5% false discovery rate at step a, so screening alone does not discriminate at this instrument size; the signature is defined by the joint model. In MVMR, ApoB (β = 0.582, P = 1.6×10⁻¹³), SBP (β = 0.658, P = 1.1×10⁻⁷) and T2D (β = 0.150, P = 1.2×10⁻⁵) each retained an independent direct effect, whereas HDL cholesterol (P = 0.31) and triglycerides (P = 0.40) collapsed. The ApoB estimate was almost identical to that obtained with the Lotta instrument (0.582 versus 0.583). Critically, the two collapsing exposures were the most strongly instrumented in the model (conditional F 64.8 and 38.7 against 37.6 for ApoB), so their loss of effect reflects causal redundancy with ApoB rather than weak-instrument bias (Table S1). Concordance of the total effect, the mediating signature and the full analytical pipeline across two instruments sharing only five of fifty-three variants, and across three outcome datasets, establishes the lean axis as a robust and reproducible finding.

## Discussion

Across the four quadrants of the adiposity–metabolic health plane, coronary risk tracked metabolic dysfunction rather than body weight: the metabolically unhealthy obese and unhealthy lean phenotypes conferred comparable harm despite occupying opposite extremes of the weight spectrum, while both metabolically healthy poles were associated with lower risk. The central finding of this study, however, lies not in the magnitude of these effects but in their architecture. Applying an identical mediator panel to each quadrant, we found that the two phenotypes at comparable coronary risk reach the disease through distinct and largely non-overlapping causal signatures: the unhealthy obese phenotype through a triglyceride–diabetes axis, and the unhealthy lean phenotype through an ApoB–blood pressure–diabetes axis. The two routes converge on T2D, where their effects are statistically indistinguishable; in every other respect the discriminating conduits are distinct. Two individuals at opposite ends of the scale, at equal risk of infarction, arrive there by different biological roads.

This bidirectional divergence reframes a body of work that has, until now, examined the pieces separately. The opposing effects of favourable and unfavourable adiposity on coronary disease were established by Martin and colleagues;^2^ the normal-weight metabolically obese phenotype linking insulin resistance to hypertension, diabetes, and coronary disease was described by Yaghootkar and colleagues a decade ago;^3^ the origin of that phenotype in limited peripheral adipose storage was demonstrated by Lotta and colleagues;^5^ and the mediation of the body-mass-index–coronary relationship through triglycerides and glycaemic traits was quantified by Xu and colleagues.^14^ Each illuminated one corner of the plane. Our contribution is to place all four quadrants within a single causal framework and to show that the risk of the lean and obese phenotypes cannot be reduced to a shared mechanism. From an endocrine perspective, these findings support the concept that cardiometabolic risk is determined not simply by the amount of adipose tissue, but by its functional capacity to safely store excess energy. Adipose tissue dysfunction, rather than adiposity per se, is therefore the common upstream abnormality linking apparently divergent body-composition phenotypes to coronary disease: the same storage failure produces risk at opposite extremes of the weight spectrum.

This work contributes on five fronts: it places all four quadrants of the adiposity–metabolic health plane within a single causal framework against a common outcome; it tests the equivalence of risk between the two high-risk phenotypes formally rather than by comparing significance; it resolves their conduits with an identical mediator panel; it reproduces the entire analytical pipeline with an independent instrument; and it distinguishes causal redundancy from weak-instrument bias using conditional instrument strength. That the obese phenotype reaches coronary disease through triglycerides and diabetes confirms prior mediation analyses of BMI,^14^. What has not been shown before is the causal architecture of the lean phenotype. Although the normal-weight metabolically obese individual has long been recognized as carrying excess coronary risk,^3,15^ the conduits of that risk had not been resolved: it was unknown whether the lean phenotype simply recapitulates the obese one at lower body weight, or whether it reaches disease by its own route. We find the latter. The lean phenotype acts through ApoB and SBP—an atherogenic, pressure-driven axis—while the triglyceride and adipokine conduits that characterize the obese phenotype carry no independent effect within it: the lean phenotype raises triglycerides more strongly than the obese one, yet once ApoB and blood pressure are accounted for they contribute nothing further to its coronary risk. The two signatures are not two intensities of one process but two distinct causal programmes, sharing only an insulin-resistance– dysglycaemia axis. This distinction is not one of degree: it means that the risk of the lean phenotype cannot be captured by the mediators of the obese one, and that the two cannot be collapsed into a single, weight-indexed model of coronary risk. To our knowledge, neither the ApoB–blood pressure architecture of the lean phenotype nor its formal dissociation from the obese architecture has previously been described.

The biological coherence of this divergence follows from the adipose expandability hypothesis.^9,10^ When the expansion capacity of subcutaneous adipose tissue is exceeded, the problem is not limited to an increase in circulating triglycerides. Abnormal fatty acid flux toward the liver and skeletal muscle develops, leading to ectopic lipid accumulation and the generation of bioactive lipid metabolites, disruption of insulin signalling, and ultimately insulin resistance and its cardiometabolic sequelae.^1216^ In the lean phenotype, in which peripheral storage capacity is constitutively limited, this lipotoxicity manifests as a lipoprotein-driven, atherogenic and hypertensive process: the genetic elevation of ApoB is the causal signature of a heightened atherogenic particle burden, consistent with the established primacy of ApoB over cholesterol mass and triglyceride content as the driver of coronary risk.^17–19^ In the obese phenotype, the same storage failure occurs against a background of expanded fat mass and manifests predominantly as hypertriglyceridaemia and dysglycaemia.^20–22^ That the two phenotypes move distinct sets of mediators indicates that these are not two intensities of one process but two distinct causal programmes converging on a shared insulin-resistance–dysglycaemia axis, ultimately culminating in overt T2D.

The robustness of the lean signature deserves emphasis. We reproduced it in its entirety with an independent instrument constructed nearly a decade later and by different methods:^7^ the same ApoB–blood pressure–diabetes signature emerged from 351 variants that share little with the original 53. Concordance of a causal signature across two independent instruments and three outcome cohorts moves this finding from a plausible association to a reproducible feature of the biology.

These findings support the functional criterion at the centre of the 2025 Commission framework, and identify a limit to its reach. Support first: the two obese quadrants differ not in degree but in direction. Adiposity with preserved metabolic function was protective (OR 0.28, 95% CI 0.19– 0.42), while adiposity with metabolic dysfunction conferred substantial risk (1.90, 1.42–2.55). The distinction between preclinical and clinical obesity therefore corresponds to a real difference in causal architecture, not to a threshold along a continuum of fat mass. The limit is this: because the diagnostic algorithm requires excess adiposity to be confirmed before organ function is assessed, a metabolically unhealthy phenotype without excess adiposity falls outside the framework entirely. Our data indicate that this phenotype carries risk indistinguishable from that of the unhealthy obese quadrant (1.86, 1.52–2.27; ratio of odds ratios 1.02, 0.72–1.46, P = 0.91), and reaches disease by a different route. Such an individual is neither preclinical nor clinical obesity, and is invisible to an algorithm that begins with the tape measure. These findings carry a direct clinical message. The lean patient who suffers a coronary event is not a statistical anomaly to be explained away, but the predictable consequence of a risk architecture that conventional weight-centred assessment does not capture. Because the atherogenic burden of this phenotype is carried by ApoB^23,24^—and, as a residual and genetically fixed component, by lipoprotein(a)^25,26^—it is precisely these measures, rather than BMI or even the standard lipid panel, that identify the lean individual at risk.^27,28^ The corollary is equally consequential: because the causal levers differ between phenotypes, a single risk-mitigation strategy cannot serve both. Risk reduction in the obese phenotype aligns with triglyceride and glycaemic control; in the lean phenotype it aligns with ApoB lowering and blood pressure management. Precision, here, means recognising that two patients at the same coronary risk require different questions asked of them. Our findings should not be interpreted as implying mutually exclusive therapeutic targets. Established cardiovascular risk factors, particularly ApoB-containing lipoproteins and blood pressure, remain clinically relevant irrespective of adiposity phenotype. Rather, our results suggest that the relative contribution of specific metabolic pathways may differ between phenotypes

The interpretation of the metabolically healthy obese quadrant requires particular care, because its genetic estimate must not be mistaken for evidence that obesity is cardioprotective—it is not, under any circumstance. The status of “metabolically healthy obesity” should be regarded as a relative and phenotype-dependent construct rather than the absence of metabolic disease. Conventional definitions may fail to capture subclinical insulin resistance, ectopic hepatic fat, or other early metabolic abnormalities. What the favourable adiposity instrument captures is not protection conferred by fat, but a greater capacity for safe lipid storage in subcutaneous depots: the ability to accommodate surplus energy without immediate metabolic decompensation. This capacity is finite and temporary. Longitudinal cohorts show that the large majority of metabolically healthy obese individuals transition to a metabolically unhealthy state over time,^29–34^ at which point they join the high-risk unhealthy obese quadrant. The genetic estimate therefore reflects a transient physiological reserve, not a stable protective state; it describes where an individual sits today, not where the trajectory leads. Within this transient window, the single conduit of residual risk we identified was lipoprotein(a): genetically raised by favourable adiposity and causally linked to coronary disease,^35,36^ yet—unlike the metabolic reserve itself— neither transient nor modifiable by lifestyle. Beyond lipoprotein(a), no circulating biomarker accounted for residual risk, locating it in the eventual exhaustion of storage capacity and the transition to metabolic decompensation rather than in a stable mediator. The leptin findings are also physiologically informative. The marked divergence in leptin between adiposity phenotypes, coupled with the absence of an independent causal effect on CAD, supports its interpretation primarily as a marker of adipose tissue mass and energy storage status rather than as a direct mediator of coronary risk in this setting.

Several limitations temper these conclusions. We restricted analyses to individuals of European ancestry, and the divergent signatures require confirmation in other populations—a priority given the disproportionate burden of the metabolically unhealthy lean phenotype in South Asian and Latin American populations.^37,38^ We examined the healthy lean quadrant as a directional consistency analysis rather than an independent discovery, and its estimates mirror the unhealthy lean quadrant by construction. We did not report point estimates of the total mediated proportion, because collinearity among conduits inflates the product-of-coefficients estimator; we established the independent contributions through conditional multivariable models instead.^39^ The multivariable models were built from the mediators that survived the exposure-to-mediator step in the same data, so their P values are conditional on that selection and are anticonservative. The replication with the mvIR instrument makes this limit explicit: at 357 variants, 24 of the 30 mediators passed a 5% false discovery rate at step a, so screening alone does not identify the signature and the discrimination rests on the multivariable model. Heterogeneity in the multivariable models was substantial (Cochran’s Q 1,177 on 305 degrees of freedom in the replication model), as expected for instruments of this size; it is balanced rather than directional, and outlier-corrected estimates remain close to the primary ones. Genetic evidence of mediation establishes a causal ordering but not a temporal sequence: it does not follow that a conduit acts early or late in the natural history of either phenotype. Several mediators are also not independent of how the instruments were defined — fasting insulin defines the lean instrument, and triglycerides, high-density lipoprotein cholesterol and sex hormone-binding globulin were among the traits used to separate the adiposity clusters — and these are reported as such rather than as findings. Effects are expressed per unit of each instrument, and because the adiposity and insulin-resistance instruments are scaled to different exposures, the similarity in magnitude between the two unhealthy quadrants should be read as concordance of risk rather than as biologically equivalent increments of exposure. Finally, MR estimates lifelong average effects and cannot capture the temporal dynamics—including phenotypic transition—that are likely central to the residual risk of the metabolically healthy obese state.

Within these bounds, our study establishes a clear and reproducible principle: convergent coronary risk can arise from divergent causal architectures. The lean and the obese do not represent one disease along a spectrum of weight, but two distinct roads to the same endpoint— and recognising which road a given patient is on is the first step toward treating the right one.

## Data Availability

All data referred to in this manuscript are publicly available. The analysis used summary-level genome-wide association statistics obtained through the IEU OpenGWAS database; accession identifiers for every exposure, mediator and outcome dataset are given in Table 3 and Table S3. No individual-level data were used. The derived genetic instruments, all result tables, the figures, and the complete analysis code are openly deposited at Zenodo under a Creative Commons Attribution 4.0 licence (DOI: 10.5281/zenodo.21682531). The deposit is a data and code archive and does not constitute prior publication.

## Acknowledgments

We thank the investigators and participants of the CARDIoGRAMplusC4D consortium, the FinnGen study, the UK Biobank, and the genome-wide association studies whose summary statistics made this work possible, as well as the IEU OpenGWAS database for curating and providing access to these data.

During the preparation of this manuscript, the authors used a generative artificial intelligence tool to improve the grammar, spelling and readability of the text. The authors reviewed and edited all output and take full responsibility for the content of this publication.

## Author contributions

J.A.S.-C. conceived and designed the study, performed the analyses, and wrote the manuscript. H.L.-M. and P.C.-C. contributed to the interpretation of the data and to the critical revision of the manuscript. E.C.-D.l.S. contributed to the interpretation of the data and to the critical revision of the manuscript. All authors reviewed and approved the final version of the manuscript.

## Non-standard Abbreviations and Acronyms

ApoB: apolipoprotein
B BMI: body mass index
CAD: coronary artery disease
FA: favourable adiposity
GWAS: genome-wide association study
IVW: inverse-variance weighted
MR: Mendelian randomization
MVMR: multivariable Mendelian randomization
mvIR: multivariate insulin resistance
SBP: systolic blood pressure
T2D: type 2 diabetes
UFA: unfavourable adiposity

## Central Illustration

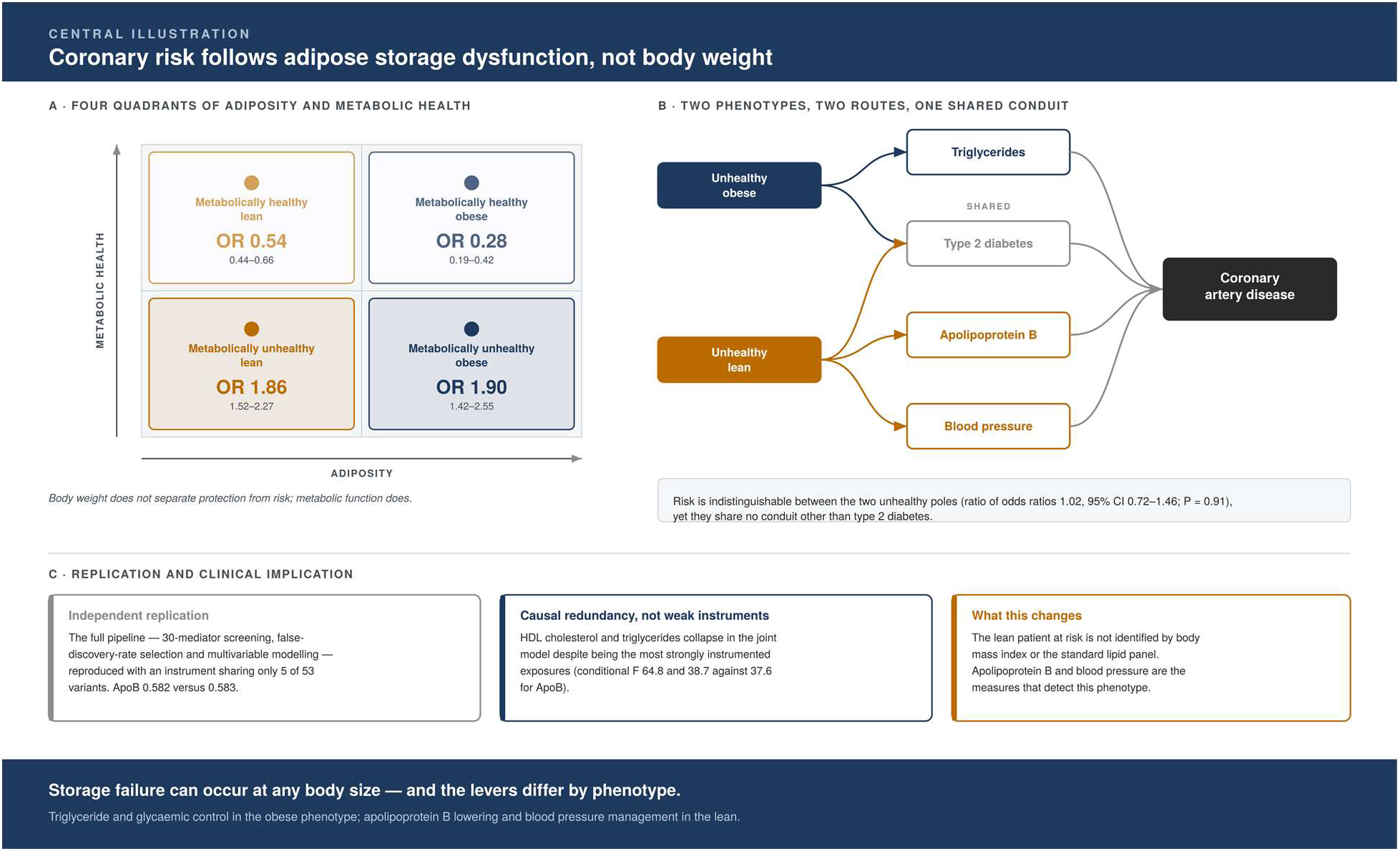

